# A lifecourse perspective on sociodemographic intersectional inequalities in physical and mental health outcomes

**DOI:** 10.64898/2026.10.23.26366317

**Authors:** Lucy Prior, George Leckie

## Abstract

Persistent inequalities in health outcomes continue to be a stubborn feature of many developed Western societies. To effectively understand health inequalities researchers must address two key criticisms: an overt focus on singular dimensions of disadvantage and a continuing need for longitudinal studies. Lifecourse intersectionality offers a framework to address these critiques through situating more nuanced and multidimensional understandings of the development of inequalities over time. We conduct a longitudinal Multilevel Analysis of Individual Heterogeneity and Discriminatory Accuracy (MAIHDA) examining both physical and mental health SF12 scores using UK Household Longitudinal Study data for adults (2009 to 2023). Forming intersections based on sex, ethnicity, generational cohort, education, and socioeconomic class, we identify a substantial degree of intersectional variation in our outcomes, and distinct differences in the development of intersectional inequalities in physical and mental health over the lifecourse. Intersectional inequalities are more pronounced for mental health compared to physical health, particularly at younger and older ages. In both cases, inequalities are additively patterned with generation and sex appearing most important to mental health and socioeconomic differences being stronger for physical health. However, our results also emphasise heterogeneity in the identified lifecourse intersectional effects, particularly in relation to how the patterning of the sociodemographic components for mental health vary by generational cohort. We therefore help reveal important complexity in the development of health inequalities in the UK and highlight the utility of longitudinal intersectional MAIHDA as a tool for quantitative intersectional researchers.

## Introduction

Many developed Western societies continue to be characterised by persistent and pervasive inequalities in health outcomes (Mackenbach, 2012; Case and Kraftman, 2024; Townsend and Davidson, 1982; Prior and Manley, 2018). A ‘social gradient’ is present, whereby those occupying positions of socioeconomic disadvantage typically experience worse health outcomes (Case and Kraftman, 2024; Costa-Font and Hernández-Quevedo, 2012; Scambler, 2012) such as lower life expectancy, poorer wellbeing and general health, as well as greater incidence of conditions such as cardiovascular disease or depression (Castelletti et al., 2024; Conrad et al., 2024; Lorant et al., 2003; ONS, 2025a). This morbidity burden places a strain on social and health services. For example, an estimated 148.9 million workdays were lost due to sickness in 2024 in the UK (ONS, 2025b). Additionally, avoidable differences in health outcomes along social, economic and environmental dimensions create a moral imperative to address such disparities to support thriving communities and a more equitable society. Therefore, understanding and addressing such social health inequalities should remain a key policy target (Ayres et al., 2024; Bibby et al., 2024; Bleich et al., 2012; GOV.UK, 2025; HM Government, 2022).

Part of the work to tackle inequalities involves firstly understanding their current state and distribution, as well as their relationship to different socioeconomic and demographic characteristics as markers of risk and exposure. To do so effectively researchers must address two key criticisms which have been levelled at studies of socioeconomic inequalities. The first is an overt focus on singular axes of disadvantage or individual ‘risk factors’ (Merlo et al., 2017), and a corresponding inattention to interactions and intersectionality (Richardson and Brown, 2016). The second is a continuing preponderance of cross-sectional studies (Jones and Rice, 2004; Van Ham and Manley, 2012), particularly in respect of multidimensional and intersectional articulations of lifecourse inequalities (Bell et al., 2024).

### Beyond singular axes of disadvantage

Regarding the first criticism, a lack of attention to multidimensionality in the factors associated with different health outcomes may come with several drawbacks. Such an approach may encourage ‘blaming the victim’ through framing inequalities as individual-level issues (Evans et al., 2018). The tendency to attach categories of exposure to individuals, without thought to the social context in which such exposures are embedded, is reflected in policy interventions which focus on addressing lifestyle factors over attempts to shift ‘upstream’ societal and economic pressures which place different groups at heightened risk (Holman et al., 2021). Additionally, singular, broad categories of identity are likely to mask important aspects of heterogeneity as individuals are enmeshed in multiple overlapping and entangled systems of oppression and marginalisation. Where there is significant heterogeneity in outcomes, relying on the ‘average’ for singular group categories could also lead to weak inferential or discriminatory accuracy in the identification of those at risk of poor health (Subramanian et al., 2018; Merlo, 2018; Merlo et al., 2017). This could feedback to policy interventions: policy could be ineffective when implemented on a more universal basis where variation around overall group averages would suggest a more targeted or proportionally tailored solution to be beneficial (Merlo et al., 2019). Overall, an overt focus on singular facets of (dis)advantage in studies of health inequalities is to the detriment of knowledge on the distinct vulnerability or resilience that may be present around the ‘on average’ story.

Intersectionality is a framework in which to situate more nuanced and multidimensional understandings of inequalities in health and other social outcomes. Intersectionality articulates the mutually constituting nature of social systems and centralises marginalised and ‘invisible’ groups to expose and critique configurations of power within social systems, both of privilege and disadvantage (Prior et al., 2025; Crenshaw, 1989; Choo and Ferree, 2010). Quantitative intersectional research relies primarily on an ‘inter-categorical’ (McCall, 2005) approach that adopts categorical definitions of identity (such as male/female or White or Black ethnicity etc.) to explore variation within and between groups and their interactions. There is a body of thinking that critiques the treatment of social categories, emphasising their inability to capture the messy realities of lived experience and highlighting their ultimately social construction (Cho et al., 2013; Choo and Ferree, 2010, McCall, 2005). However, such identity markers continue as societal signifiers of persistent hierarchies of power relations (MacKinnon, 2013). Therefore, bringing forward intersectional perspectives remains a promising avenue for advancing the study of health inequalities (Holman and Walker, 2021).

Emerging techniques for analysing intersectional dynamics in health outcomes, such as Multilevel Analysis of Individual Heterogeneity and Discriminatory Accuracy (MAIHDA) (Evans et al., 2024a; Evans et al., 2020; Evans et al., 2018; Merlo, 2018) may be particularly appropriate here. This approach has been growing in popularity within epidemiology and the social sciences (Prior et al., 2025; Persmark et al., 2019; Fisk et al., 2018; Moreno-Agostino et al., 2024; Evans et al., 2023; Hernández-Yumar et al., 2018; Keller et al., 2023; Alvarez and Evans, 2021). It facilitates the operationalisation of intersectional thinking within quantitative studies in a manner more aligned with the tenets of intersectionality, situating intersectional identities as contexts within which individuals are embedded and socialised. This implementation helps avoid ‘blaming the victim’ and maintains an awareness for the structural power hierarchies which shape the identity categories we utilise within quantitative research (Evans et al., 2020). Methods such as intersectional MAIHDA offer ways to explore and identify diverse patterns of intersectional dynamics as part of toolkit of approaches to comprehend health inequalities in a multidimensional manner.

### Lifecourse perspectives on intersectionality

Cross-sectional studies still predominate the literature on health inequalities, necessarily limiting the capacity for research to uncover insight into the development of inequalities through time. Fortunately, there has been a considerable drive towards lifecourse research on health, with lifecourse epidemiology and conceptual models for health pathways emerging since the 1990s (Blane et al., 2007; Kuh et al., 2003; Wadsworth, 1997), as well as a growing body of cohort and other longitudinal datasets by which to explore trajectories of health relationships (Blaxter, 1986; CLOSER, 2025). However, extensions of lifecourse research to consider intersectional entanglements (or conversely of intersectional literature to deliberate the nature and impact of ageing and lifecourse transitions) are less prevalent (Holman and Walker, 2021).

The exploration of health trajectories through an intersectional lens will provide insights beyond those of static cross-sectional studies. An intersectional framework on longitudinal and lifecourse health studies would forefront conceptualisations of the heterogeneous nature of ageing, including differing trajectories, transitions and life events (Ferrer et al., 2017; Holman and Walker, 2021). Differing lifecourse hypotheses are posited for how health inequalities will evolve with time, such as ageing as leveller, persistent inequality, cumulative inequality, sensitive/critical periods, and chains of risk (Ferraro and Farmer, 1996; Kuh et al., 2003). These may be present to a greater or lesser degree for different intersections of status/identity and may also vary by health outcome measured. For instance, evidence from older adults in the United States has suggested that Black women may experience an accelerated increase in functional limitations compared with Black men, White or Mexican American individuals, who show more constant disparities through middle to old age according to the ‘persistent inequality hypothesis’ (Warner and Brown, 2011). Brown et al., (2016) examined intersections of age, race/ethnicity, gender and socioeconomic status and revealed that the gap between higher status Black and White individuals increased with age as the highly educated Black grouping experienced heightened declines in their health, in line with a cumulative disadvantage perspective.

Combining intersectionality theory with lifecourse studies would help situate dynamic understandings of identity within diverse contexts of space, place and time and how changing social systems shape developing health inequalities along intersectional lines. For example, Ferrer et al. (2017) highlighted the interplay of ageing, caring labour and immigration with racialized and gendered inequalities through a case study of a Filipino community in Canada. Brown (2018) noted the potential role of racialised immigrant integration processes that serve to marginalise and stigmatise Black and Mexican American immigrants, with deleterious consequences for their health trajectories through middle to later life. Therefore, the synthesis of lifecourse intersectionality will provide insights into the production, maintenance or breakdown of health. In turn, these new insights could inform policy decisions on where, when and for whom to target interventions for tackling inequalities. However, space remains for further research utilising a lifecourse intersectional framework, particularly for work which considers the whole lifecourse, rather than focusing on later-life ageing. Recent extensions to the MAIHDA methodology have shown how this promising technique can be combined with longitudinal analyses (Bell et al., 2024) and are particularly pertinent to bringing forward lifecourse understandings of the development of sociodemographic intersectional health inequalities.

### Heterogeneity across different outcomes

An appreciation for complexity is vitally important both to providing nuanced empirical understandings of health inequalities and to intersectional thinking. One aspect of heterogeneity in health inequalities is how intersectional variation may present differently across different health dimensions. Gendered and culturally shaped attitudes towards physical and mental health conditions could manifest in intersectional differences between these outcomes. For example, a stigma can still exist around mental illness and this may be more apparent in reporting patterns for men where there are gendered expectations of stoic masculinity (Gorski, 2010). Additionally, physical health and mental health conditions can often show comorbidity (Rai et al., 2016; Wienand et al., 2024). An intersectional framework could help reveal multiple marginalised communities at risk of social, environmental and other stressors that may contribute to an enhanced burden of multiple conditions, for instance through a chronic weathering on the body (Seeman et al., 2014; Forde et al., 2019).

Furthermore, introducing temporality to the discussion could reveal extra dimensions of heterogeneity between physical and mental health. For example, societal and economic conditions such as a recession may have a more marked effect on mental health for certain groups, such as young adults poised to enter the job market (Alam and Bose, 2022). In general, older persons and generations may see declining physical health but intersections of privileged characteristics may offset some of this decline through maintenance of positive mental wellbeing whilst disadvantaged groups suffer from their history of socialised discrimination and marginalisation (Gilmore-Bykovskyi et al., 2021; Sadana et al., 2016). Through examination of both physical and mental health trajectories under an intersectional lens we can uncover these important aspects for understanding the development of inequalities.

### This study

This paper will help address the dearth of longitudinal intersectional studies and thus tackle the two key critiques of health inequalities studies discussed above. We will use a longitudinal MAIHDA analysis to examine physical and mental health trends over the lifecourse for individuals in the UK. The overall aim of the study is to examine sociodemographic intersectional variation in these two interrelated dimensions of health and quality of life, and how they develop over time.

### Data

This study utilises the UK Household Longitudinal Survey (UKHLS) dataset. UKHLS is a panel survey of households, where all individuals within a selected household are surveyed. We will be using 14 waves of data running from 2009 to 2023. Due to the survey design, we can assess a broad range of ages (16 to 96) through a compilation of different generational cohorts.

The two outcome health measures are the SF12 mental and physical health metrics. These are short-form versions of the widely used SF36 health measures and are quality of life measures that express the degree to which these dimensions of health impact on everyday life (Ware et al., 1995). The physical health metric covers: physical functioning, role-physical, bodily pain, and general health. The mental health metric covers: vitality, social functioning, role-emotional, and mental health. Both are continuous scores ranging from 0 to 100 where higher numbers are indicative of higher functioning or better health, and the two scores can be meaningfully compared (Ware et al., 2002).

UKHLS also features a variety of social and demographic measures that we can use to define intersectional groups. For this study we focus on facets of identity that proxy for key systems of marginalisation and discrimination, as well as axes which are routinely linked to health differentials. These are sex (male, female), ethnicity (White, Black, Asian, Mixed, Other), highest educational qualification achieved (degree or other higher, A level/GCSE/other qualifications, no qualifications) and socioeconomic class (management and professional, intermediate, routine). Additionally, as we are assessing intersectional variation over time we need to consider the temporal identities of our study population. Therefore, generational cohort will also be included as we can reasonably expect there to be differences in health outcomes across cohorts as they progress through life under different economic, political, social and technological climates. The generational cohorts are: Silent Generation (birth years 1945 and earlier), Baby Boomers (born 1946 to 1963), Generation X (born 1964 to 1979), Generation Y (born 1980 to 1994) and Generation Z (born 1995 or later), following the example of Bell et al., (2024).

Recorded changes in ethnicity or birth year (used to define generational cohort) were treated as erroneous and cases were dropped on this basis. In the case of education, we chose to use the highest achieved qualification (a time invariant variable) as this aligns more with the educational status concept we are trying to capture. To operationalise the concept of socioeconomic class we utilise the highest modal category across observations (also time invariant) as this is thought to better reflect overall social status, rather than potentially short-term changes in class based on occupational positions. Changes in recorded sex may reflect genuine experiences and highlight the potential marginalisation of transgender communities. However, as there were so few instances of this in our population (<0.1%) we chose to drop these cases and continue with a static formulation of sex identity. Age will be included in the fixed part to model trajectories in mental and physical functioning and will be centred around age 50, as a ‘middle-aged’ individual. A descriptive summary of the outcomes, age and intersectional components is shown in Table 1, with further details on missingness, category changes over time and the distribution of occasions provided in the Supplementary Materials. Our final sample of those with complete non-missing information across all our sociodemographic variables was 298,911 occasions, nested in 50,622 individuals, within 372 strata.

**Table 1.** Descriptive statistics.

| **Variable** | **Descriptive Statistics** |
| --- | --- |
| Physical health | Mean = 51.99, SD = 8.97 |
| Mental health | Mean = 48.94, SD = 9.92 |
| Age | Mean = 43.15, SD = 15.46, Range: 16-96 |
| Sex | Male: 45.2%<br>Female: 54.8% |
| Ethnicity | White: 83.2%<br>Mixed: 1.9%<br>Asian: 9.9%<br>Black: 4.2%<br>Other: 0.9% |
| Education | No qualifications: 4.0%<br>Other qualifications: 6.7%<br>GCSE or equivalent: 18.3%<br>A level or equivalent: 21.6%<br>Other higher qualifications: 13/9%<br>Degree: 35.5% |
| Socioeconomic class | Routine: 25.7%<br>Intermediate: 21.2%<br>Management and Professional: 53.2% |
| Generation | Silent Generation: 3.0%<br>Baby Boomers: 29.4%<br>Generation X: 34.5%<br>Generation Y: 24.7%<br>Generation Z: 8.4% |
| Strata | N = 372 |
| Individuals | N = 50,622 |
| Occasions | N = 298,911 |

## Methods

This study relies on the MAIHDA methodology which offers many advantages over traditional approaches to studying intersectional relationships (notably the inclusion of interaction terms between two or more axes of identity in regression analyses). These include: easily and parsimoniously accommodating several different dimensions of status: directly quantifying the degree to which intersectional grouping account for total variation in outcomes; reducing the risk of overinterpreting strata predictions that may have arisen by chance; and enabling the analysis of variation within and between strata, affording appreciation for multiple strands of heterogeneity (Prior et al., 2025; Bell et al., 2019; Leckie et al., 2025; Merlo, 2018).

We employ a longitudinal intersectional MAIHDA in this study. Under the longitudinal design the two-level formulation of individuals nested within their intersectional strata is extended to three-levels: occasions nested within individuals within strata. This set-up is possible as we are considering our intersectional strata as unchanging through time for this exploratory examination of health inequalities.

For a cross-sectional intersectional MAIHDA two models are typically run. The first is a variance components model without any covariates which assesses the overall degree of variation present at the level of your intersectional strata (Evans et al., 2024a). The second model then adds the components of the intersectional strata as covariates. This model allows researchers to analyse the degree to which the intersectional variation is composed of main effects versus interaction effects (additive versus multiplicative effects) with any remaining intersectional level variation being the result of two or higher-way interactions between the intersectional components.

For the longitudinal formulation as used in this study, these models are extended by the inclusion of age terms in the fixed part of the model and through allowing these age effects to vary at higher levels. This enables us to examine trajectories of health through time and how these trajectories vary between individuals and by intersectional groups. The first set of models can be written as follows:

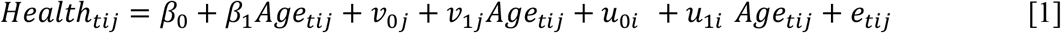

This first model features a linear (*β*_1_) effect of age, which will we centre around age 50 so that the intercept *β*_0_ provides an estimate of the overall average physical or mental health for ‘middle-aged’ individuals. For simplicity we only present the linear age effect, but the model could also be extended by further polynomial terms. We allow the intercepts to vary at the stratum level (level-3) (*v*_0j_) as well as allowing the age slope (the linear portion) to vary across intersectional strata (*v*_1j_). Thus, we are modelling stratum specific mean differences in the average level of health (the intercepts) but also how health develops and changes over time (the age slopes/trajectories). Similarly, we allow the intercepts and age trajectories to vary at the individual level (level-2) through the *u*_0ij_ and *u*_1ij_ terms, capturing further heterogeneity within intersectional strata. At the lowest level (level-1, occasions) the residual *e*_tij_ summarises remaining variation not accounted for by higher levels, that is within individuals, between occasions.

The second set of models can be written as follows:

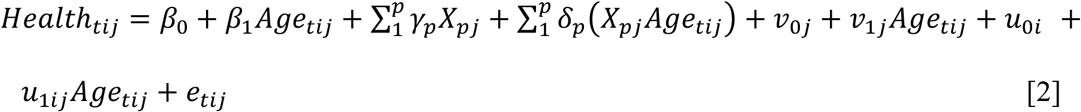

This model set extends the first through the inclusion of the main effects (γ_p_) of the intersectional strata components in the fixed portion of the model, here represented by *X*_pj_ for *p* stratum-level dummies. Additionally, as we are interested in health trajectories, we additionally include interactions effects (*δ*_p_) of all strata components with age.

Examination of the variance is an important element to the analysis. The stratum- and individual-level random effects of models 1 and 2 fall under the typical normality assumptions of a multilevel model. As these are random slope models the variance at the higher levels (strata and individuals) are expressed as a variance function. This function shows how the variance, which can be considered here a measure of the extent of inequalities across or within intersections, changes with age.

An important statistic is the Variance Partitioning Coefficient (VPC). A stratum-level VPC assesses the proportion of overall variation that is situated between intersections, providing quantification for the importance of the chosen intersectional identities to outcome inequalities. In this longitudinal study, the stratum-level VPCs will vary as a function with age. Alongside the overall stratum VPC, we can also calculate separate VPC’s for the intercepts and slopes. The intercept VPC will calculate the extent to which intersectional differences account for overall mean differences in health for the reference 50-year-old person. The Slope VPC explains the degree that differences in the changes in physical or mental health over time are intersectional in nature.

The motivation for fitting the second model is to determine the extent to which additive or multiplicative effects between the intersectional components explain overall intersectional variation. This helps distinguish whether intersectional effects are operating on a more universal basis across different stratum axes or whether there is heterogeneity in how intersectional inequalities operate. The relative importance of additive effects (the sociodemographic main effects) is given by the Proportion Change in Variance (PCV), which will vary with age. Additionally, intercept and slope specific PCVs will be calculated. The intercept PCV will demonstrate how overall mean intersectional differences are explained by additive rather than interactive effects. The slope PCV will do the same but in reference to differences in the health slopes.

In this study we will be examining two outcomes: physical and mental health. Separate sets of the models above will be run for physical and mental health, before comparisons are made between the outcomes. Further detail on the methodology, including algebraic expressions of the variance components, functions and calculation of VPCs and PCVs is provided in the Supplementary Materials.

## Results

### Variance components

Tables 2 and 3 provide the model results predicting the physical health (Table 2) and mental health (Table 3) SF12 scores. Intersectional variation is higher for the mental health score where our strata account for approximately a quarter of variation in the mean scores for our middle-aged individuals (intercept VPC: 26.9%) and age trajectories (slope VPC: 22.6%). In contrast, for physical health the proportion of intercept (mean score) and slope (age trajectory) variation explained by intersectional strata is lower at 10.2% and 6.1% respectively. This shows that our intersectional effects are likely to be much more influential to the overall mental health and functioning of individuals and how this develops over time than they will be to physical health. The results do indicate a large share of variability in outcomes lies within each intersection, between individuals and/or across occasions. Thus, the differences between individuals in any given stratum exceed the mean differences across strata, see Supplementary Figures S1 and S2 for VPC and variance plots including all three levels.

**Table 2.** Model results predicting physical health score by age (Model 1), and age plus the stratum components (Model 2).

| *Outcome = Physical Health* |  | Model 1 |  | Model 2 |  |  |  |
| --- | --- | --- | --- | --- | --- | --- | --- |
|  |  | Est. | S.E | Est. | S.E | Est. | S.E |
| *Fixed effects* |  |  |  |  |  |  |  |
| Intercept |  | 50.567 | 0.175 | 51.583 | 0.250 |  |  |
| Age-50 |  | -0.266 | 0.007 | -0.275 | 0.013 |  |  |
| Age-50<sup>2</sup> |  | -0.006 | 0.000 | -0.005 | 0.000 |  |  |
| Age-50<sup>3</sup> |  | -0.00004 | 0.000 | 0.00001 | 0.000 |  |  |
|  |  |  |  | *Main effects* |  | *Interactions* |  |
| Sex (ref. = Male) | Female |  |  | -1.177 | 0.166 | -0.025 | 0.007 |
| Ethnicity<br>(ref. = White) | Mixed |  |  | -1.084 | 0.421 | -0.026 | 0.018 |
|  | Asian |  |  | -2.591 | 0.226 | -0.028 | 0.010 |
|  | Black |  |  | -1.121 | 0.277 | -0.018 | 0.013 |
|  | Other |  |  | -2.069 | 0.483 | -0.038 | 0.024 |
| Highest Qualification<br>(ref. = Degree level) | No qualifications |  |  | -1.975 | 0.279 | -0.023 | 0.014 |
|  | A level/GCSE/Other |  |  | -1.068 | 0.182 | -0.015 | 0.008 |
| Socioeconomic Class<br>(ref. = Professional) | Routine |  |  | 1.971 | 0.206 | 0.041 | 0.009 |
|  | Intermediate |  |  | 0.899 | 0.206 | 0.029 | 0.009 |
| Generation<br>(ref. = Generation X) | Silent Generation |  |  | 6.883 | 0.591 | -0.108 | 0.034 |
|  | Baby Boomers |  |  | 1.002 | 0.208 | 0.038 | 0.016 |
|  | Generation Y |  |  | -1.484 | 0.320 | -0.059 | 0.018 |
|  | Generation Z |  |  | -1.240 | 0.721 | -0.076 | 0.030 |
| *Random effects* |  |  |  |  |  |  |  |
| Level 3: Stratum | Variance: Intercepts | 4.966 | 0.678 | 0.396 | 0.126 |  |  |
|  | Covariance: Intercepts-Slopes | 0.106 | 0.020 | 0.011 | 0.005 |  |  |
|  | Variance: Slopes | 0.003 | 0.001 | 0.001 | 0.000 |  |  |
| Level 2: Individual | Variance: Intercepts | 43.857 | 0.487 | 43.850 | 0.486 |  |  |
|  | Covariance: Intercepts-Slopes | 1.130 | 0.021 | 1.128 | 0.021 |  |  |
|  | Variance: Slopes | 0.047 | 0.001 | 0.047 | 0.001 |  |  |
| Level 1: Occasion | Variance | 34.132 | 0.104 | 34.130 | 0.104 |  |  |
| VPC Intercepts |  | 10.2% |  | 0.9% |  |  |  |
| VPC Slopes |  | 6.1% |  | 1.5% |  |  |  |
| PCV Intercepts |  |  |  | 92.0% |  |  |  |
| PCV Slopes |  |  |  | 76.5% |  |  |  |

**Table 3.** Model results predicting mental health score by age (Model 1), and age plus stratum components (Model 2).

| *Outcome = Mental health* |  | Model 1 |  | Model 2 |  |  |  |
| --- | --- | --- | --- | --- | --- | --- | --- |
|  |  | Est. | S.E | Est. | S.E | Est. | S.E |
| *Fixed effects* |  |  |  |  |  |  |  |
| Intercept |  | 47.575 | 0.259 | 48.735 | 0.236 |  |  |
| Age-gm |  | 0.043 | 0.012 | -0.111 | 0.016 |  |  |
| Age-gm<sup>2</sup> |  | 0.001 | 0.000 | 0.002 | 0.000 |  |  |
| Age-gm<sup>3</sup> |  | -0.0002 | 0.000 | -0.0001 | 0.000 |  |  |
|  |  |  |  | *Main effects* |  | *Interactions* |  |
| Sex (ref. = Male) | Female |  |  | -1.921 | 0.159 | 0.039 | 0.009 |
| Ethnicity<br>(ref. = White) | Mixed |  |  | -0.722 | 0.422 | 0.027 | 0.021 |
|  | Asian |  |  | -0.184 | 0.218 | -0.023 | 0.013 |
|  | Black |  |  | 1.245 | 0.269 | 0.023 | 0.016 |
|  | Other |  |  | 0.530 | 0.488 | 0.047 | 0.028 |
| Highest Qualification<br>(ref. = Degree level) | No qualifications |  |  | 0.070 | 0.273 | 0.004 | 0.017 |
|  | A level/GCSE/other |  |  | 0.033 | 0.174 | 0.016 | 0.010 |
| Socioeconomic Class<br>(ref. = Professional) | Routine |  |  | 0.515 | 0.197 | -0.006 | 0.012 |
|  | Intermediate |  |  | 0.373 | 0.198 | -0.002 | 0.012 |
| Generation<br>(ref. = Generation X) | Silent Generation |  |  | 7.575 | 0.668 | 0.018 | 0.041 |
|  | Baby Boomers |  |  | 2.615 | 0.196 | 0.078 | 0.020 |
|  | Generation Y |  |  | -4.793 | 0.344 | 0.003 | 0.022 |
|  | Generation Z |  |  | -9.687 | 0.861 | 0.023 | 0.037 |
| *Random effects* |  |  |  |  |  |  |  |
| Level 3: Stratum | Variance: Intercepts | 14.455 | 2.033 | 0.288 | 0.143 |  |  |
|  | Covariance: Intercepts-Slopes | 0.028 | 0.051 | 0.011 | 0.007 |  |  |
|  | Variance: Slopes | 0.012 | 0.002 | 0.002 | 0.000 |  |  |
| Level 2: Individual | Variance: Intercepts | 39.290 | 0.489 | 39.402 | 0.489 |  |  |
|  | Covariance: Intercepts-Slopes | 0.352 | 0.022 | 0.345 | 0.022 |  |  |
|  | Variance: Slopes | 0.041 | 0.002 | 0.040 | 0.002 |  |  |
| Level 1: Occasion | Variance | 50.324 | 0.153 | 50.342 | 0.153 |  |  |
| VPC Intercepts |  | 26.9% | 0.7% |  |  |  |  |
| VPC Slopes |  | 22.6% | 4.0% |  |  |  |  |
| PCV Intercepts |  |  | 98.0% |  |  |  |  |
| PCV Slopes |  |  | 86.2% |  |  |  |  |

Plotting the stratum VPC function by age from the unadjusted model further demonstrates the differences between outcomes (Figure 1). The variance function plot is provided in Supplementary Figure S3. The overall higher proportion of stratum-level variation for mental health in comparison to physical health is evident. Figure 1 also reveals that the proportion of overall variation situated between strata is higher at both younger and older ages for mental health. In contrast, we see that younger ages (around 20 to 30 years old) show the lowest proportions of stratum-level variation for physical health with this increasing as persons transition through middle age (30 to 60) before plateauing for older ages.

**Figure 1.**
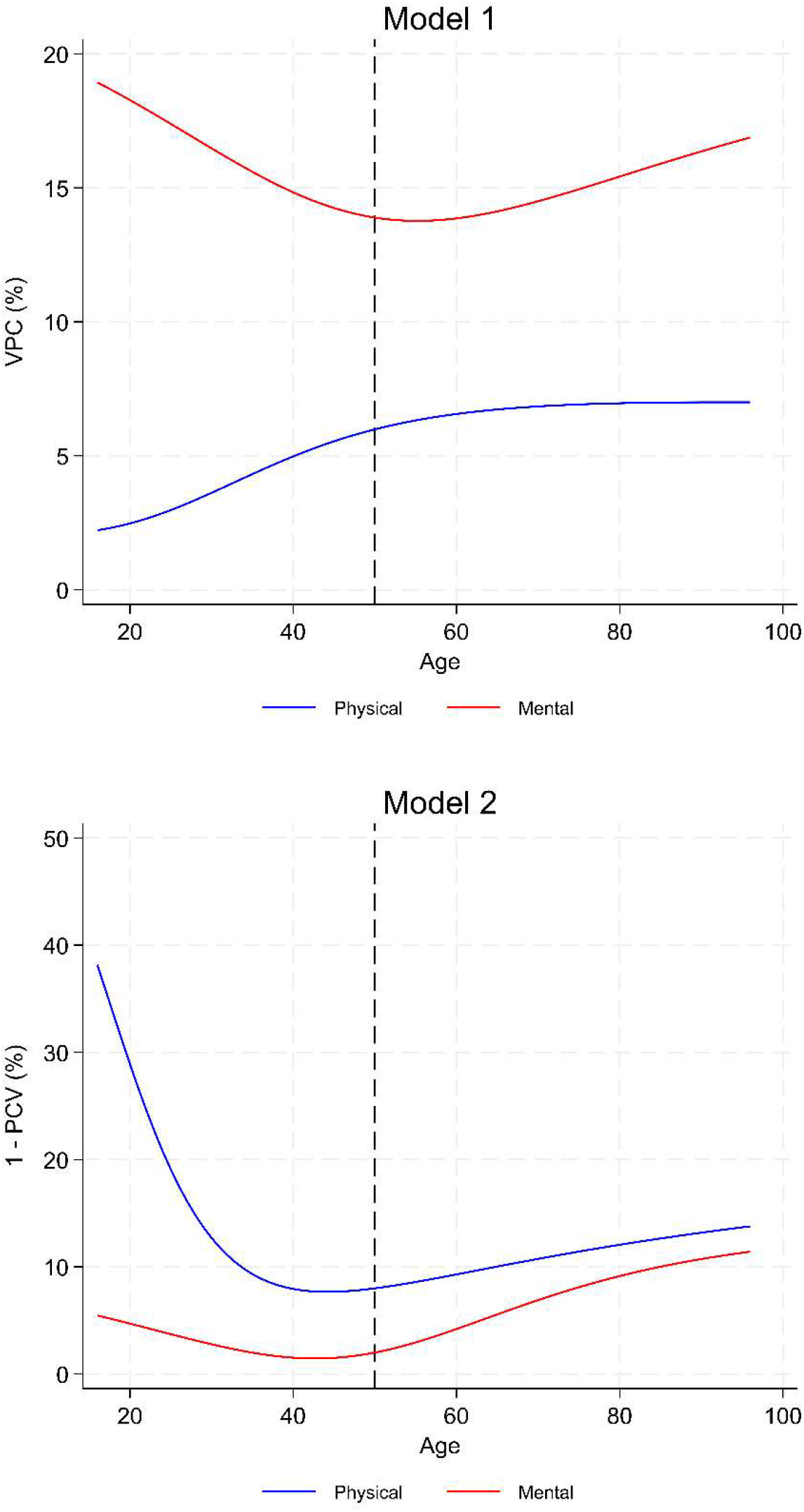
Stratum-level VPC function for Model 1 (top) and percentage non-additive stratum effects (1-PCV) for Model 2 (bottom) for physical and mental health.

Model 2, where we include the stratum defining components and interactions of these with age in the fixed portion of the model, allows us to evaluate the degree to which additive effects versus multiplicative effects account for variation in the mean health and age trajectories. The majority of the intersectional effects we identify are composed of additive effects. For physical health our model additions explain 92.0% of the intercept variance and 76.5% of the slope variance. The PCVs for mental health are higher at 98.0% and 86.2% for the intercepts and slopes respectively. Figure 1 additionally shows the percentage of variability across intersections which is non-additive (that is due to interactions) by age. It is apparent that interaction or multiplicative effects are likely to be more prominent for physical health, particularly so at younger ages where for example around 30% of variation is non-additive at age 20. Mental health sees a slight increase in the proportion of intersectional variation which is non-additive from around 60 years of age, reaching around 10% non-additive effects. Therefore, whilst the intersectional variation we identify in Model 1 is acting on a largely universal basis, with little variation across the other axes of strata identities, there is a suggestion of interactive intersectional effects and that these may vary in importance over the lifecourse.

### Predicted stratum effects

Figure 2 shows the predicted stratum effects from Model 2 (adjusted for age, stratum components and age by component interactions) coloured by sex. The difference in the overall trajectories of physical and mental health is stark. For physical health the intersectional strata show a clear downward trajectory throughout the lifecourse where the decline accelerates as individuals approach later life. In contrast, mental health shows an overall slightly positive trend over the lifecourse, but the generational effects are prominent in defining clusters of strata and show varied trends (Supplementary Figure S4 shows the generational clusters). For example, the youngest generations (Generations Z and Y) show generally declining mental health through time with strata defined by the youngest generation showing consistently worse mental health scores than Generation Y at comparable ages. Where we can compare across cohorts at the same age points there is a tendency for older generations to show better mental health than their younger generation counterparts. Generation X also shows a declining trend, but this is shallower than for Generation Y or Z. The strata group defined by the Baby Boomer cohort (ages from around 50 to 75) show flatter trajectories through time, suggesting that this group consistently maintains their mental health functioning for the ages monitored. Finally, the strata containing those of the Silent Generation start the survey period with relatively good mental health functioning, but this group then declines on an accelerating curve as they enter very old age (80 plus years).

**Figure 2.**
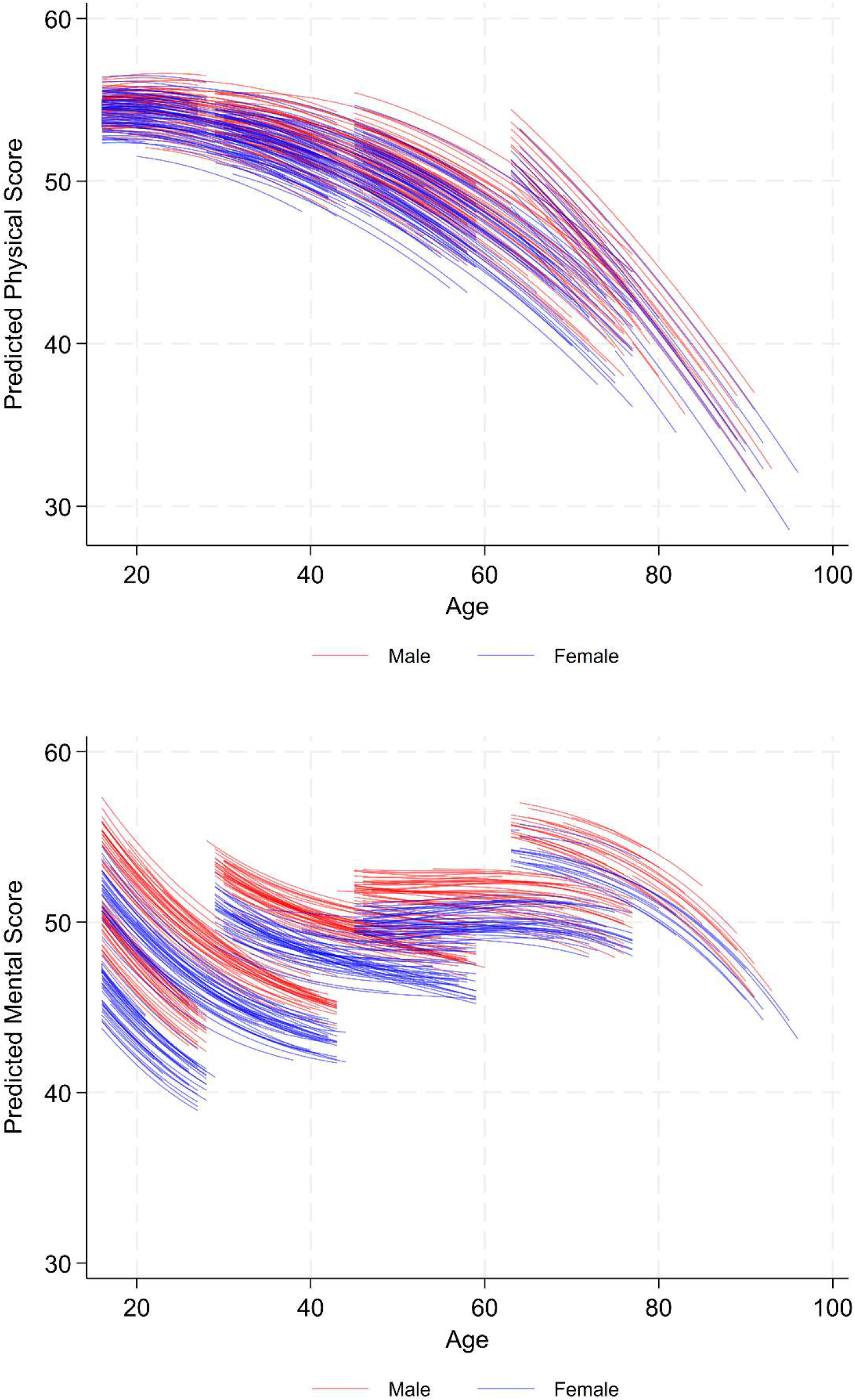
Predicted physical and mental health from Model 2 coloured by sex

Figure 2 also highlights there is a more evident sex effect for mental than physical health whereby the scores for females are consistently worse within the generational cohort groupings. In comparison this tendency is less clear for physical health where the lines for males and females across different strata groups overlap considerably along the age spectrum. We see this trend reflecting in the additive main effects where the gap between females and males for the middle-aged reference person is −1.921 in mental health score compared with −1.177 in physical health score.

The predicted stratum effects coloured by ethnicity are shown in Figure 3 (plots coloured by the remaining stratum components (generation, highest qualification, highest socioeconomic class) are provided in the Supplementary Materials). Here again we can identify differences between physical and mental health in how the effect of ethnicity plays out between strata. For physical health there is a tendency for groups containing White individuals to have generally better physical functioning than stratum defined by ethnic minorities, especially those with Asian identities. This can be seen as a reflection of the dominant main effects, where we see a trend for minority ethnicities to have worse physical health scores than White individuals, ranging from −1.084 for Mixed ethnicities, to −2.591 for Asian individuals for the reference 50-year-old. In contrast, for mental health the ethnicity patterning shows distinct clusters, where each ethnicity tends to appear in two distinct bands. This corresponds to combinations of ethnicity with sex, highlighting the strength of the sex effect where males consistently show better mental health than females.

**Figure 3.**
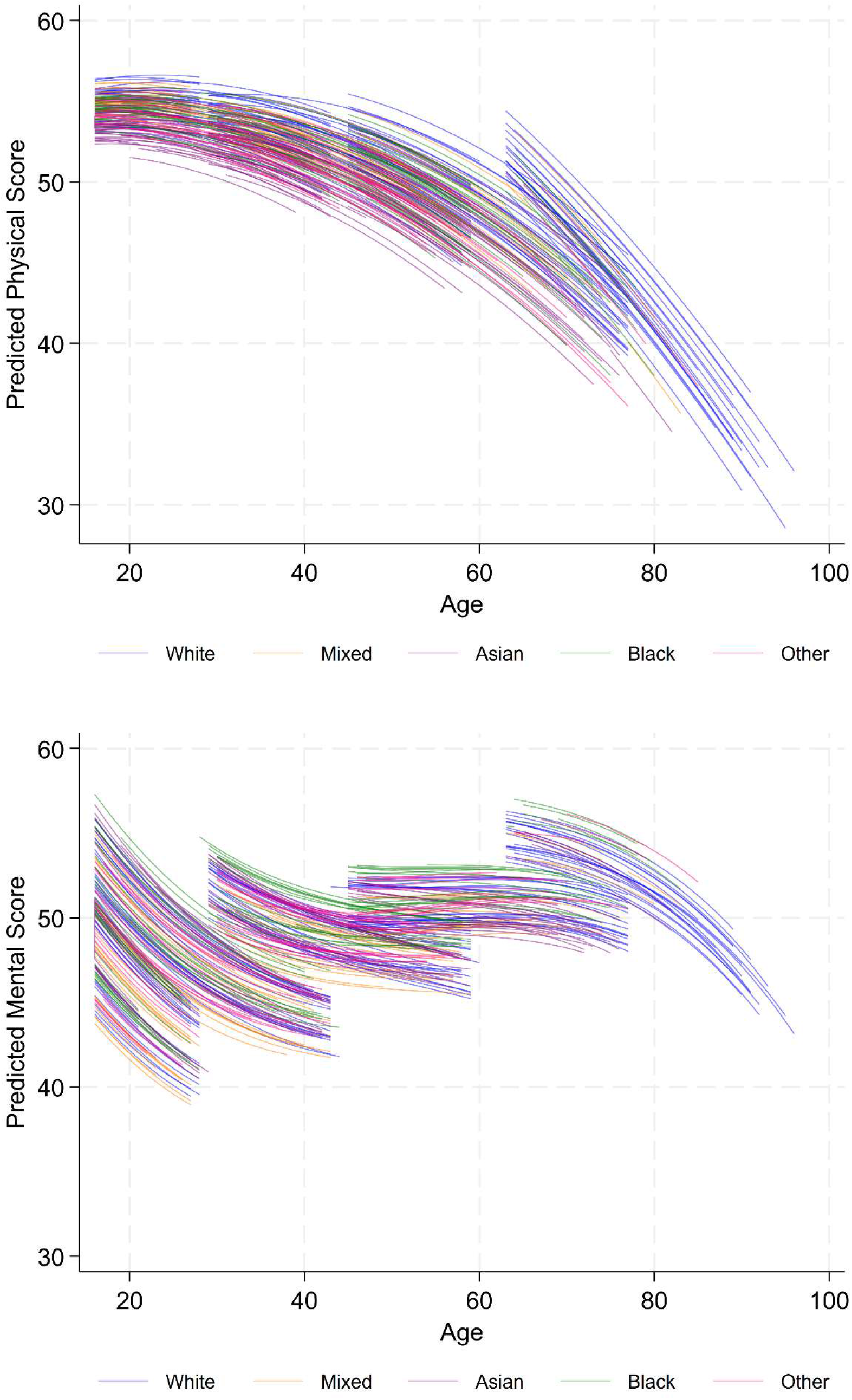
Predicted physical and mental health from Model 2 coloured by ethnicity

Examining the ethnicity by sex strata groupings in isolation (Supplementary Figures S7 to S10) reveals that those groups defined by Black individuals score higher for their mental health than their White strata counterparts. In comparison this pattern is reversed when looking at Mixed ethnicity strata, which tend to score lower than White groupings within the sex bandings. Strata composed by Asian individuals show a more mixed picture where generation also appears to matter. For the youngest cohorts (Generations Z and Y), the Asian males and Asian females typically have better mental health scores than the respective male and female strata composed of White individuals. For Generation X strata the Asian and White groupings are overlapping (still maintaining the overall sex difference) and then for Baby Boomers the two ethnicities begin to diverge so that White strata start to show better mental health. This patterning for Baby Boomers would align with a cumulative disadvantage hypothesis where the more marginalised groups show accelerated declines in health. There is clearly heterogeneity in how intersectional inequalities may be expected to play out for different cohorts and through the lifecourse. Of note for both physical and mental health is that most strata present for the oldest generational cohort (Silent Generation) are defined by White individuals. Supplementary Figure S11 examines the degree to which our strata our fully populated shown by generation, sex and ethnicity. It highlights the stark underrepresentation for minority ethnicities for the Silent Generation cohort, particularly so for females. This could be a true representation of the composition of the Silent Generation, potentially due to lower levels of immigration in the pre-war period. However, there may also be an aspect of differential selection into the study, or it could relate to intersectionally patterned attrition or survivorship.

For the socioeconomic components of the intersectional strata (highest educational qualification and socioeconomic class, Supplementary Figures S5 and S6) we see some evidence of a gradient by status for physical health. The highest status strata (those with the highest qualifications and professional or managerial positions) tend to maintain better health scores throughout the lifecourse. Whereas, for mental health there is no clear patterning by status. These trends are reflective of the additive main effects we identify where the gradients along socioeconomic dimensions for physical health are more pronounced and statistically significant. For example, for a reference middle-aged person, those with no qualifications are expected to have physical functioning that is −1.975 points lower than those with a degree level qualification, whilst for those with A level/GCSE or other qualifications this difference is −1.068.

### Correlations

The overall correlation between the intersectional stratum intercepts (overall mean health scores) for physical and mental health is 0.36 in Model 1. Thus, intersections with generally worse physical health tend to also have worse mental health but only moderately so. The correlation between the stratum slopes (age trajectories) is much weaker at just −0.06, meaning there is a very little consistency between the two dimensions of health in how they develop over time. However, it is important to consider how the overall correlation between the stratum effects for physical and mental health may vary by age. Figure 4 demonstrates how the correlation varies as we consider 20-year age slices from 20 to 80 years old. It shows that the correlation increases as we consider older individuals, from −0.17 for 20-year-olds to 0.76 for adults in their 80s. Therefore, we expect physical and mental health outcomes to become more aligned with each other as people age.

**Figure 4.**
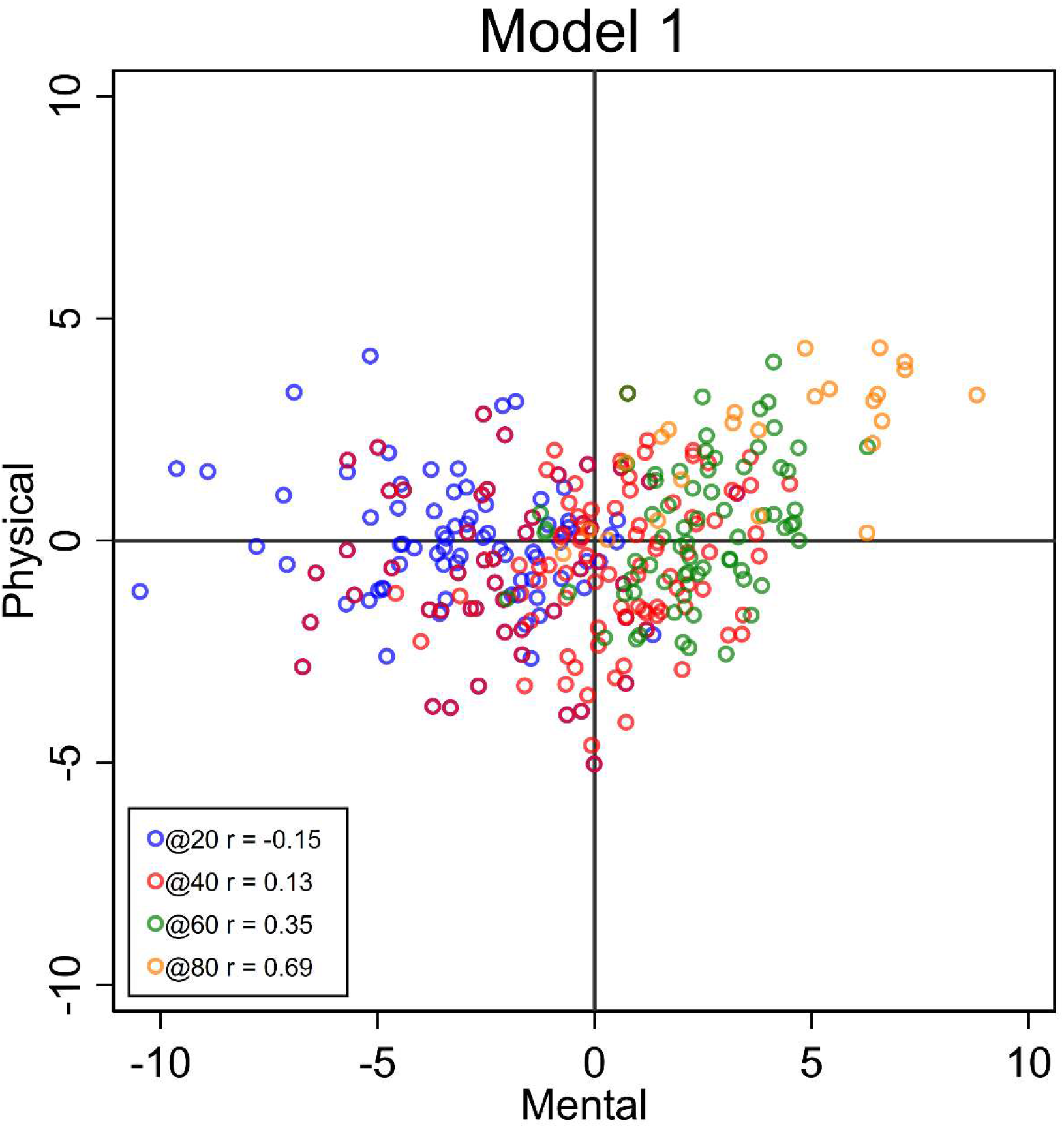
Scatterplot and correlation between the overall intersectional stratum effects for physical and mental health for ages 20, 40, 60 and 80.

Additionally, we can consider those strata which show particularly divergent effects between their physical and mental health outcomes. Here we will focus on the intercept correlations (evaluated for the age-centred 50-year-old) from Model 1, as they dominate the overall stratum effects, and we consider outlying or divergent strata to be those where the difference between the physical and mental health score is 2 or more times the standard deviation of the average difference between the two outcomes. This equates to a difference of around 6.10 points or more between intercepts. Taking these criteria, we see that most divergent strata are from the youngest generations (Generation Z (11 out of 17, 65%) and Generation Y (4 out of 17, 24%) and these are showing worse mental health than physical. Additionally, we see around half (9 out of 17, 53%) of the divergent strata feature females of either White or Mixed ethnicities. Therefore, we see many divergent cases where White or Mixed ethnicity females from the youngest generations are showing particularly worse mental health in comparison with their physical health.

## Discussion

This study used a longitudinal MAIHDA analysis to explore intersectional variation and trajectories in physical and mental health functioning scores for adults in the UK. We found significant and substantial variation in individual outcomes between our intersectional strata. This showcases the continuing importance of intersectionality as a means to avoid the ‘tyranny of averages’ and the inefficient provision of interventions based on group averages of singular disadvantage metrics (Merlo et al., 2017; Merlo et al., 2019). For instance, intersectional inequalities accounted for 27% of variation in mental health scores for middle-aged individuals and 10% for physical health. By the grading scale suggested by Fisk et al. (2018) for social epidemiological applications, this would qualify our results as “good” to “very good” in terms of the potency of our intersectional strata. In contrast, Bell et al., (2024), in their study of General Health Questionnaire (GHQ) responses (a mental health metric), reported that only 9% of the outcome variation for similarly middle-aged individuals (48.13 years). This contrast may in part be due to differences between our two mental health measures, with the SF-12 potentially covering a broader view of quality of life status and with a wider range of potential values (0-36 for GHQ compared with 0-100 for SF-12). Therefore, our strata constitute a potentially important social context by which to delineate (dis)advantaged groups in terms of their health outcomes and it is likely worthwhile for policymakers to have greater consideration for multiple dimensions of identity in aiming to tackle inequalities.

It is clear from our results that the expression of intersectional inequalities is highly heterogeneous, with no single inequality model applicable for all strata or at all ages. We found that the importance of intersectional inequalities to physical health was less potent at younger ages, but more consistent through the rest of the lifecourse, showing some agreement with a persistent inequality hypothesis. In contrast, the proportion of variation in mental health attributable to our intersectional strata increased for the oldest ages. Widening inequalities into old age could align with a cumulative disadvantage hypothesis (Brown et al., 2016), whereby those in already advantaged positions are able to accumulate greater resources or socioeconomic capital to mitigate health risks, whilst already disadvantaged groups suffer an exacerbation of risks and deleterious exposures as they age. We found that intersectional inequalities in mental health may also be especially pronounced among the younger aged participants. This could reflect a particular vulnerability, or sensitive period, at this life stage (Rod et al., 2025). Key transitions, such as those away from family environments, or into the labour market could represent important moments where intersectional differentials in social and resource capital could impact mental resilience.

The complexity of our results demonstrates the useful knowledge that can be gained from lifecourse intersectionality research using MAIHDA. It can highlight periods of heightened vulnerability for disadvantaged groups to aid with tackling inequalities. The increased inequalities at older ages (from around 60 years old) suggests that working to achieve healthy ageing for all should be a target for policymakers, particularly considering ageing populations in the UK and around the world (ONS, 2018; Sadana et al., 2016). Additionally, our comparison between physical and mental health showcased how the strata predictions can be used to identify divergent cases which may be prudent for further investigation, especially as ill health on one dimension may negatively impact on the other. For example, it appears that some intersectional groups (White females) from the youngest generations may be suffering particularly in their mental health in comparison to physical health. Addressing the mental wellbeing of young adults appears prudent as the time between 18 and 29 is now considered an important developmental period (Arnett et al., 2014). Poor mental health during this time may not only have knock-on effects for a greater burden of later mental health, as well as physical illness through comorbidities (McCloughen et al., 2012; Wienand et al., 2024), but impede successful life achievements in terms of education, relationships and occupational life which are then detrimental to later inequalities (Gustavson et al., 2018).

The MAIHDA approach also revealed that most of the intersectional effects operate on an additive basis, meaning that, for example, inequalities according to sex were operating similarly across different categories of ethnicity or socioeconomic status. Whilst the conceptual language of intersectionality prioritises the terminology of ‘interaction(s)’, the finding of dominant additive effects does not discount the importance of intersectionality to our outcomes (Evans et al., 2024b). Furthermore, our longitudinal analysis revealed that non-additive effects may vary in prominence depending on an individual’s age. Therefore, it is vitally important to incorporate time as a further dimension of heterogeneity in studies of how health inequalities develop and are maintained. In other words, the results here encourage the use of more bespoke or precision-targeted interventions, such as proportionate universalism (Marmot, 2010), to improve physical and mental health outcomes.

Overall, it is evident that longitudinal MAIHDA techniques offer a promising avenue to expand lifecourse intersectionality research. This study did not extend into a deeper study of the potential mechanisms behind any identified inequalities, offering instead an initial mapping to highlight where critical timepoints and marginalised groups may exist in overall physical and mental health functioning. Opportunities for further analysis also exist in ways to push the MAIHDA methodology forward. For example, here we considered two outcomes but ran separate models as we found the combination of random slopes with bivariate models too computationally burdensome for this dataset. However, extensions to bivariate or multivariate models (Prior et al., 2021) could be possible and would provide opportunities for more direct comparisons across outcomes in terms of the operation of intersectional effects. Similarly, the incorporation of lagged dependent variables could be uses to assess the relevant importance of state vs. trait effects including how this also potentially differs by intersectional identities (Steele, 2008). In this study we considered our intersectional strata as fixed entities, focusing on how health developed over the lifecourse for those singularly defined groupings. However, adjustments to the MAIHDA multilevel set-up to allow shifts in social belonging over time, analogous to students changing schools for example (Heck et al., 2022), would allow more nuanced understandings of intersectional inequalities. It could reveal groups whose social mobility is stymied for example. Researchers should therefore continue to progress the work of lifecourse intersectionality through these promising techniques.

## Data Availability

All data referred to are available online at http://doi.org/10.5255/UKDA-SN-6614-22

## Acknowledgements

Understanding Society is an initiative funded by the Economic and Social Research Council and various Government Departments, with scientific leadership by the Institute for Social and Economic Research, University of Essex, and survey delivery by the National Centre for Social Research (NatCen) and Verian (formerly Kantar Public). The research data are distributed by the UK Data Service. The COVID-19 study (2020-2021) was funded by the Economic and Social Research Council and the Health Foundation. Serology testing was funded by the COVID-19 Longitudinal Health and Wealth – National Core Study. Fieldwork for the web survey was carried out by Ipsos MORI and for the telephone survey by Kantar.

