## Supplementary Materials for "A lifecourse perspective on sociodemographic intersectional inequalities in physical and mental health outcomes"

In the longitudinal MAIHDA models analysed in this study the variance at level 3 (stratum, denoted by *v*) and level 2 (individual, denoted by *u*) is a function of age, whilst the occasion-level variance (denoted by *e*) is constant. The variance functions are as follows.

$$\text{Level3 variance} = \sigma_{v0}^2 + 2\sigma_{v0}\ Age_{tij} + \sigma_{v1}^2 Age_{tij}^2$$

[1]

$$\text{Level2 variance} = \sigma_{u0}^2 + 2\sigma_{u0}\ Age_{tij} + \sigma_{u1}^2 Age_{tij}^2$$

[2]

$$\text{Level1 variance} = \sigma_e^2$$

[3]

The stratum-level variance function is therefore calculated as the intercept variance ($\sigma_{v0}^2$), plus two times the intercept-slope covariance ($2\sigma_{v0}$ ) multiplied by age, plus the slope variance ($\sigma_{v1}^2$) multiplied by age$^2$. Intersectional variation is thus a quadratic function of age.

The stratum-level and individual-level variance are assumed to be independent and multivariate normally distributed with zero mean vectors and unstructured covariance matrices. The occasion-level variance is assumed independent and normally distributed.

$$\begin{pmatrix} v_{0j[n]} \\ v_{1j[n]} \end{pmatrix} \sim \mathrm{N}\left\{\begin{pmatrix} 0 \\ 0 \end{pmatrix}, \begin{pmatrix} \sigma_{v0[n]}^2 & \\ \sigma_{v01[n]} & \sigma_{v1[n]}^2 \end{pmatrix}\right\}$$

$$\begin{pmatrix} u_{0j[n]} \\ u_{1j[n]} \end{pmatrix} \sim \mathrm{N}\left\{\begin{pmatrix} 0 \\ 0 \end{pmatrix}, \begin{pmatrix} \sigma_{u0[n]}^2 & \\ \sigma_{u01[n]} & \sigma_{u1[n]}^2 \end{pmatrix}\right\}$$

$$e_{tij[n]} \sim \mathrm{N}\left(0, \sigma_{e[n]}^2\right)$$

[4]

To calculate the overall Variance Partitioning Coefficient (VPC) the following equation [5] is used. To assess the VPC as a percentage, showcasing the proportion of overall variation that lies between strata, the outcome of this equation is simply multiplied by 100. As the Level3 and Level2 variances are a function of age (as seen above) the overall VPC also varies with age.

$$\text{Overall VPC} = \frac{Level3\ variance}{Level3\ variance\ +\ Level2variance\ +\ Level1\ variance}$$

[5]

This study also makes use of the separate intercept and slope VPCs, as in equation [6].

$$\text{Intercept VPC} = \frac{\sigma_{v0}^2}{\sigma_{v0}^2 + \sigma_{u0}^2}$$

$$Slope\ VPC = \frac{\sigma_{v1}^2}{\sigma_{v1}^2 + \sigma_{u1}^2}$$

[6]

The intercept VPC is dependent on how age is centred. We chose to centre age around a ‘middle-aged’ person at age 50. Therefore, our intercept VPC calculated the extent of intersectional inequalities, as a proportion of all individual difference, for 50-year-old persons. However, the slope VPC is unaffected by centring choices and conveys the extent to which intersectional groups explain between-individual differences in the slopes of the age trajectories.

As with the VPC we can also calculate an overall Proportion Change in Variance (PCV) (equation [7]) and separate intercept and slope PCVs (equation [8]). The overall PCV provides the proportion (or percentage when multiplied by 100) of stratum-level variance that is accounted for by additive effects. Therefore, 1-PCV is a useful statistic to summarise the proportion of non-additive of interactive effects. The overall PCV is calculate as in equation [7] where the subscripts represent the model number.

$$Overall\ PCV = \frac{Level3\ variance_{[1]} - Level3\ variance_{[2]}}{Level3\ variance_{[1]}}$$

[7]

The intercept PCV informs on the extent to which additive effects of the intersectional components explain overall average variability between intersectional groups for the middle-aged person (or wherever age has been centred). The slope PCV tells us the extent to which differences in age trajectories between intersectional strata are explained by the singular interactions of the intersectional variable components with age.

$$Intercept\ PCV = \frac{\sigma^2_{v0[1]} - \sigma^2_{v0[2]}}{\sigma^2_{v0[1]}}$$

$$Slope\ PCV = \frac{\sigma^2_{v1[1]} - \sigma^2_{v1[2]}}{\sigma^2_{v1[1]}}$$

[8]

**Supplementary Tables**

***Table S1. Degree of missing data in the physical and mental health outcomes, as well as age and the sociodemographic components for the sample with outcome data***

| | Total | Missing | % |
|---|---|---|---|
| Physical health | 568,946 | 72,248 | 12.70 |
| Mental health | 568,946 | 72,248 | 12.70 |
| Age | 496,968 | 92 | 0.02 |
| Birth year (for generation) | 496,698 | 266 | 0.05 |
| Sex/gender | 496,698 | 8 | 0.00 |
| Ethnicity | 496,698 | 89,409 | 18.00 |
| Education | 496,698 | 4,893 | 0.99 |
| SES | 496,698 | 134,239 | 27.03 |
| Total by age and sociodemographic variables | 496,698 | 197,787 | 39.82 |

***Table S2. Total number and percentage of state changes (moving between variable categories) made for the sociodemographic intersectional components***

| | Number of state changes | % |
|---|---|---|
| Birth year (for generation) | 3,561 | 0.72 |
| Sex/gender | 456 | 0.09 |
| Ethnicity | 70,937 | 14.28 |
| Education | 62,295 | 12.54 |
| SES | 249,648 | 50.26 |

***Table S3. Distribution of number of occasions individuals are present in the dataset***

| Number of occasions | Frequency | Percent |
|---|---|---|
| 1 | 11,189 | 3.74 |
| 2 | 9,856 | 3.3 |
| 3 | 11,520 | 3.85 |
| 4 | 25,892 | 8.66 |
| 5 | 13,185 | 4.41 |
| 6 | 13,038 | 4.36 |
| 7 | 13,461 | 4.5 |
| 8 | 14,112 | 4.72 |
| 9 | 14,517 | 4.86 |
| 10 | 16,310 | 5.46 |
| 11 | 19,470 | 6.51 |
| 12 | 25,620 | 8.57 |
| 13 | 41,665 | 13.94 |
| 14 | 69,076 | 23.11 |

*Table S4. Strata showing the largest difference between physical and mental health intercepts from Model 1*

| Stratum ID | Sex | Ethnicity | Highest Qualification | Socioeconomic Class | Generation | Stratum Intercepts | | Difference |
|---|---|---|---|---|---|---|---|---|
| | | | | | | Physical | Mental | |
| 6 | Male | White | A level/GCSE/Other | Professional | Silent | 3.24 | 9.92 | -6.68 |
| 1 | Male | White | No qual. | Routine | Silent | 0.68 | 6.82 | -6.14 |
| 300 | Male | White | A level/GCSE/Other | Routine | Generation Z | 1.93 | -4.54 | 6.47 |
| 264 | Female | White | Degree/Other higher | Routine | Generation Y | 2.17 | -4.31 | 6.48 |
| 336 | Female | White | No qual. | Professional | Generation Z | 0.65 | -5.86 | 6.51 |
| 272 | Female | Mixed | Degree/Other higher | Intermediate | Generation Y | 1.56 | -5.15 | 6.71 |
| 345 | Female | Mixed | A level/GCSE/Other | Intermediate | Generation Z | 1.54 | -5.92 | 7.46 |
| 268 | Female | Mixed | A level/GCSE/Other | Routine | Generation Y | 2.01 | -5.57 | 7.58 |
| 349 | Female | Mixed | Degree/Other higher | Professional | Generation Z | -0.04 | -7.63 | 7.59 |
| 291 | Female | Other | A level/GCSE/Other | Routine | Generation Y | 1.16 | -6.53 | 7.69 |
| 339 | Female | White | A level/GCSE/Other | Professional | Generation Z | -1.49 | -9.41 | 7.92 |
| 303 | Male | White | Degree/Other higher | Routine | Generation Z | 4.05 | -4.12 | 8.17 |
| 337 | Female | White | A level/GCSE/Other | Routine | Generation Z | 0.95 | -7.23 | 8.18 |
| 370 | Female | Other | Degree/Other higher | Routine | Generation Z | 1.49 | -7.31 | 8.80 |
| 302 | Male | White | A level/GCSE/Other | Professional | Generation Z | 1.20 | -8.36 | 9.56 |
| 305 | Male | White | Degree/Other higher | Professional | Generation Z | 3.50 | -7.02 | 10.52 |
| 342 | Female | White | Degree/Other higher | Professional | Generation Z | 1.78 | -8.98 | 10.76 |

**Supplementary Figures**

***Figure S1. VPC and variance plots for Model 1 showing stratum, individual and occasion levels for physical health***

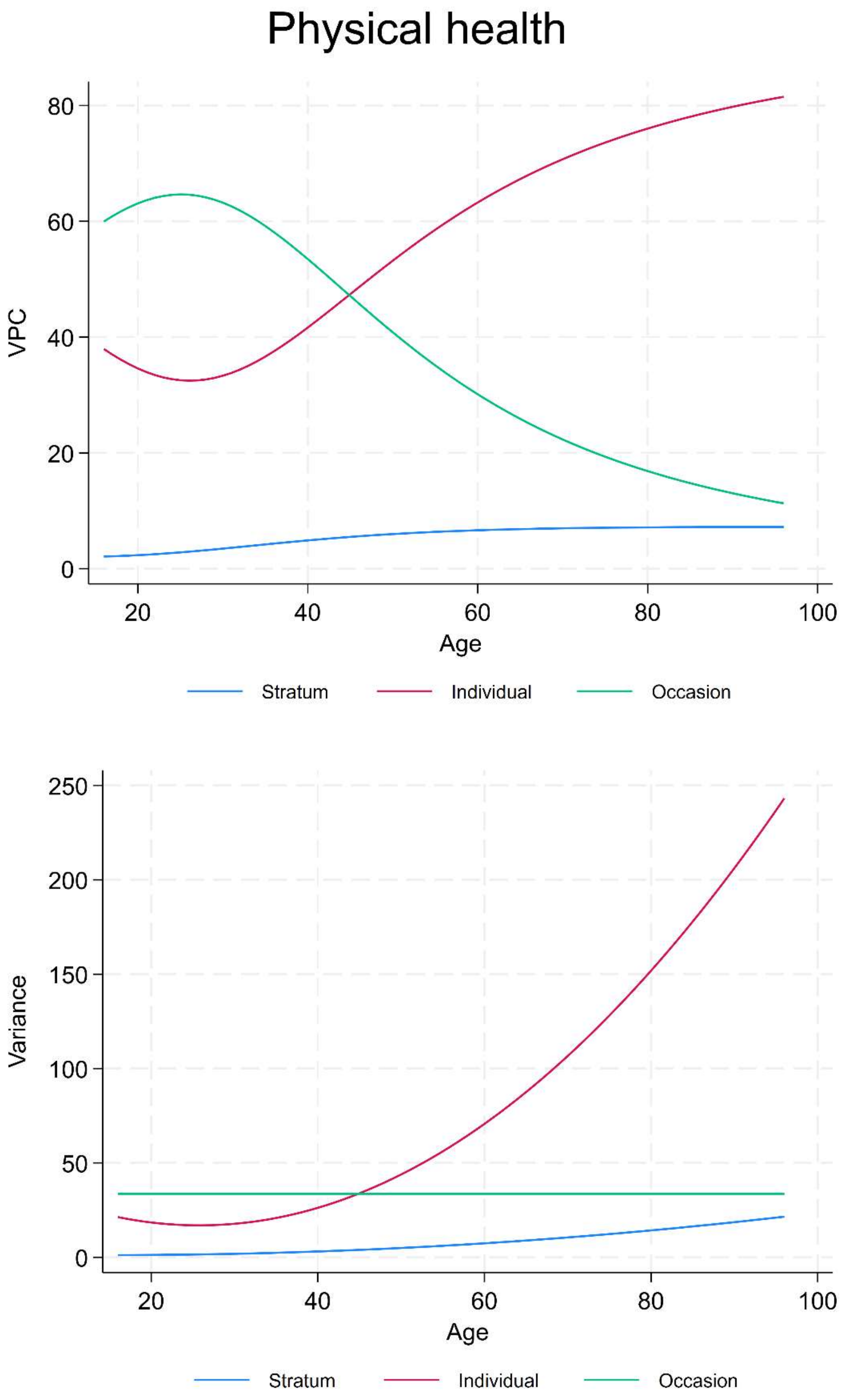

***Figure S2. VPC and variance plots for Model 1 showing stratum, individual and occasion levels for mental health***

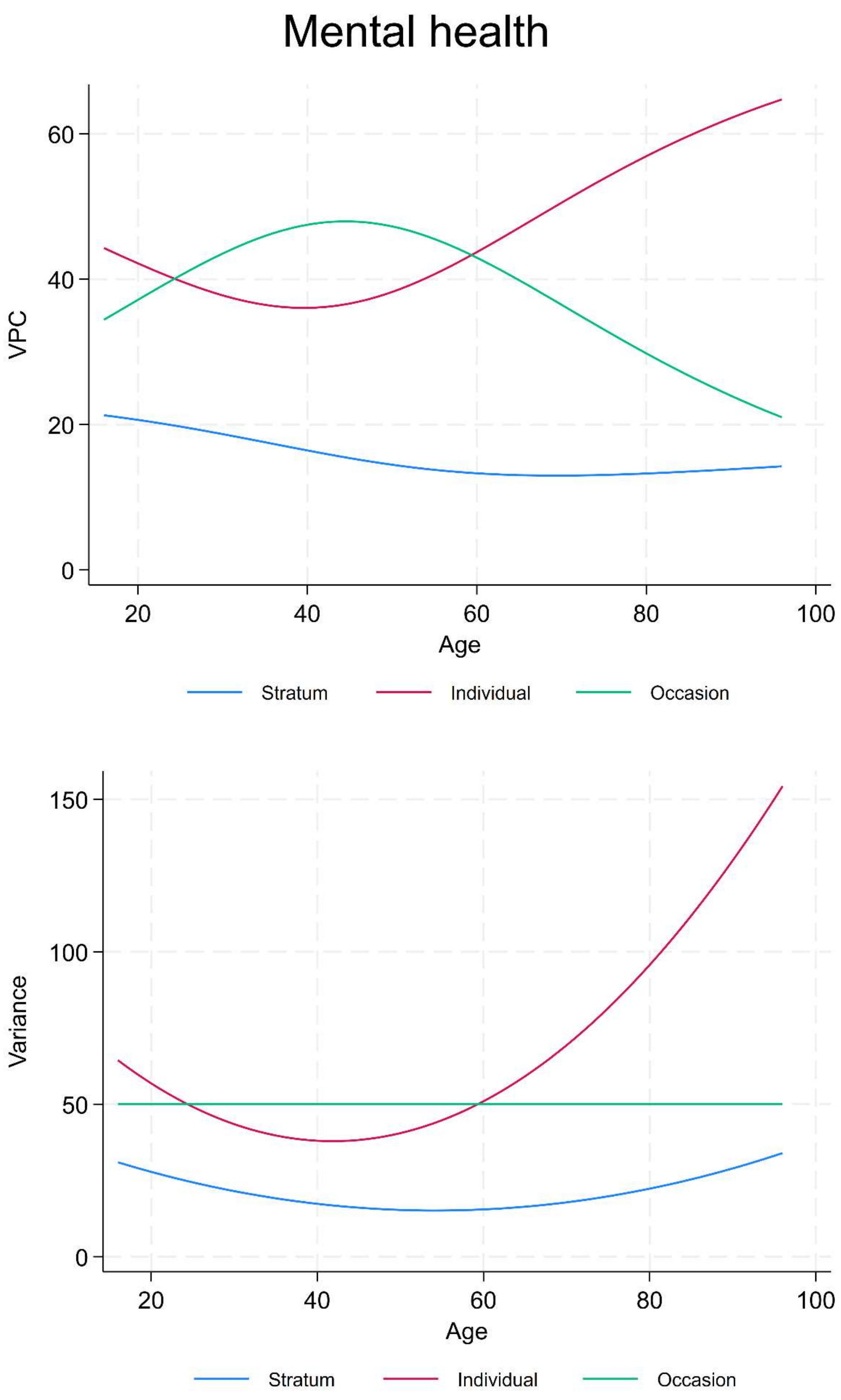

*Figure S3. Stratum-level variance function for physical and mental health outcomes for Model 1 (age) and Model 2 (age plus stratum components)*

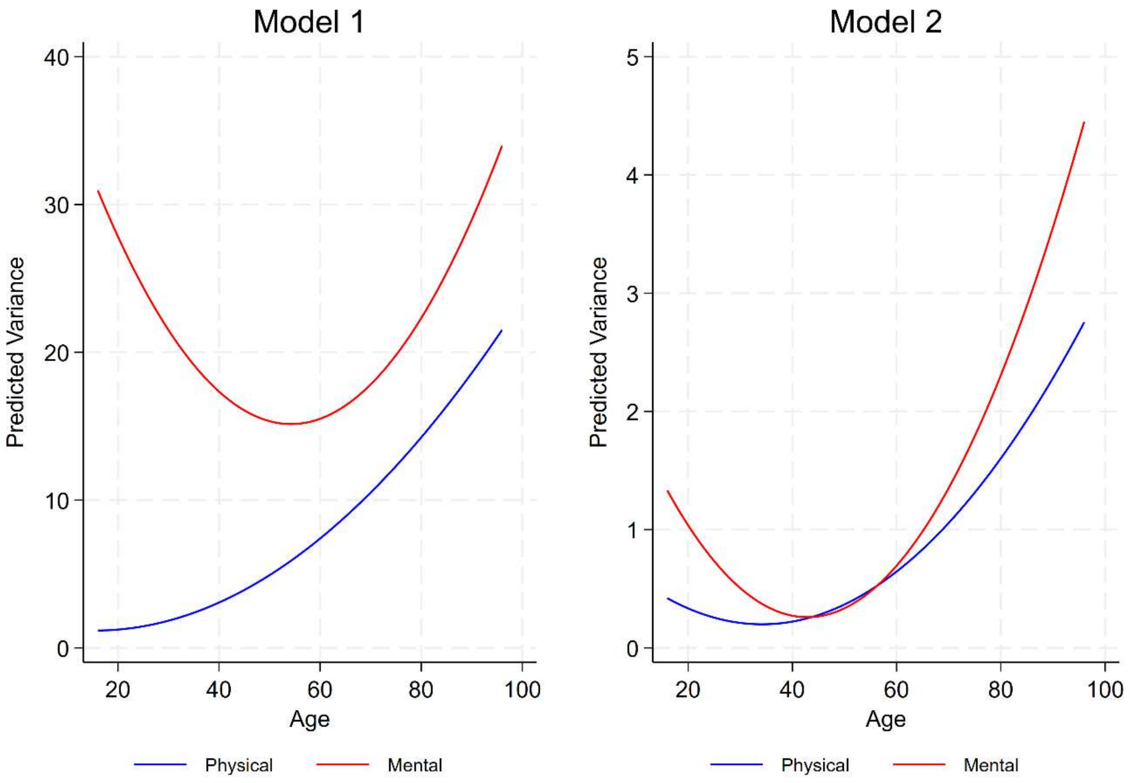

***Figure S4. Predicted physical and mental health from Model 2 coloured by generation***

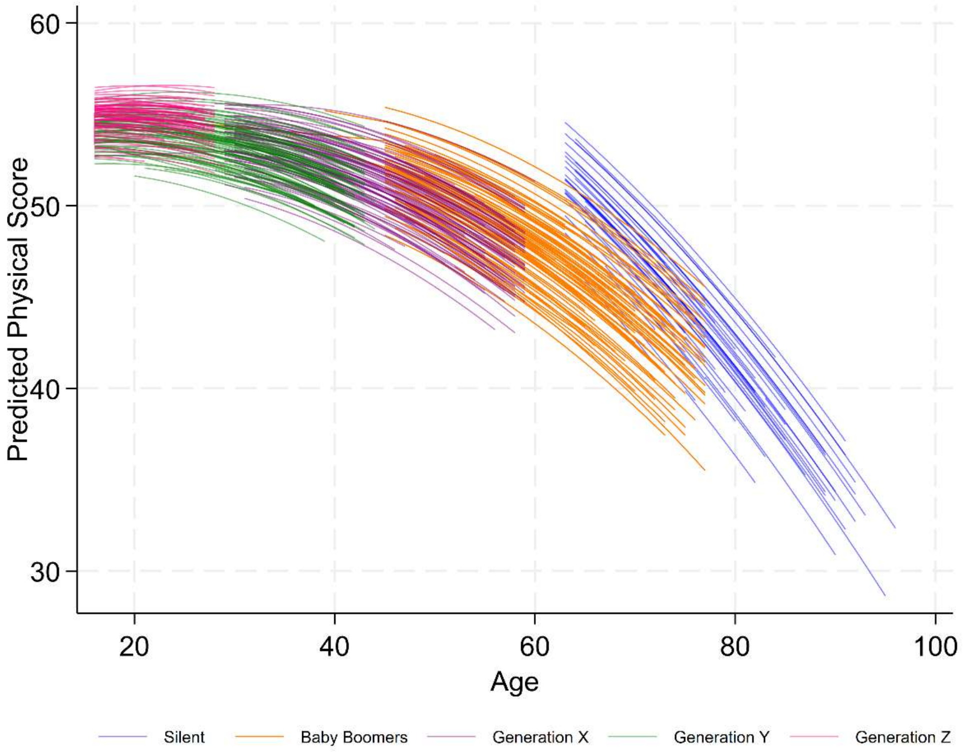


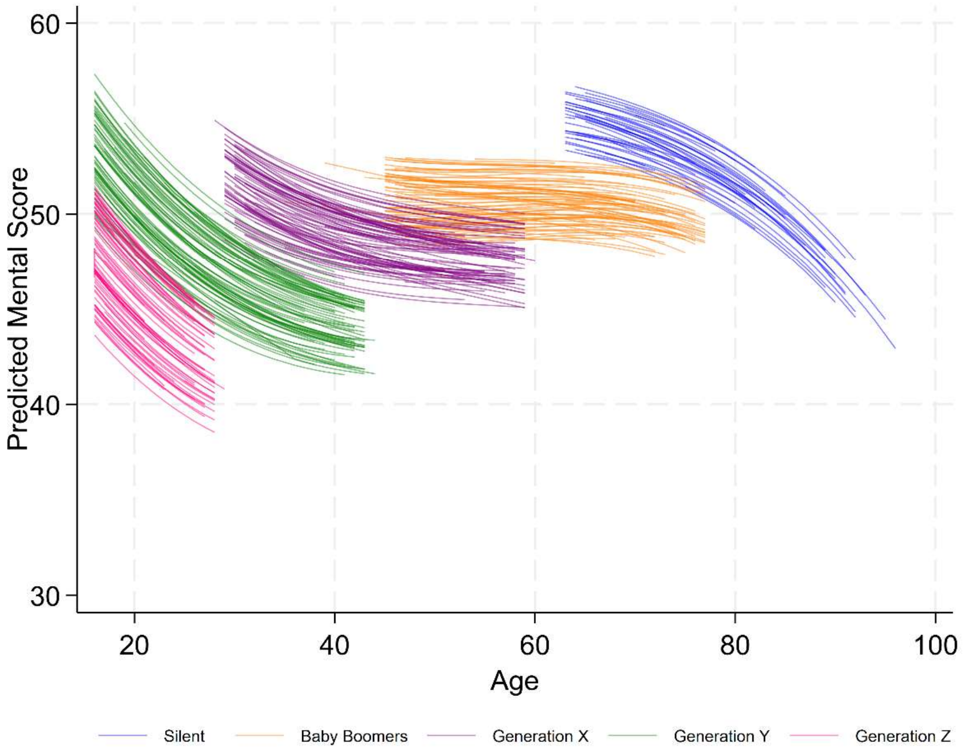

*Figure S5. Predicted physical and mental health from Model 2 coloured by highest qualification*

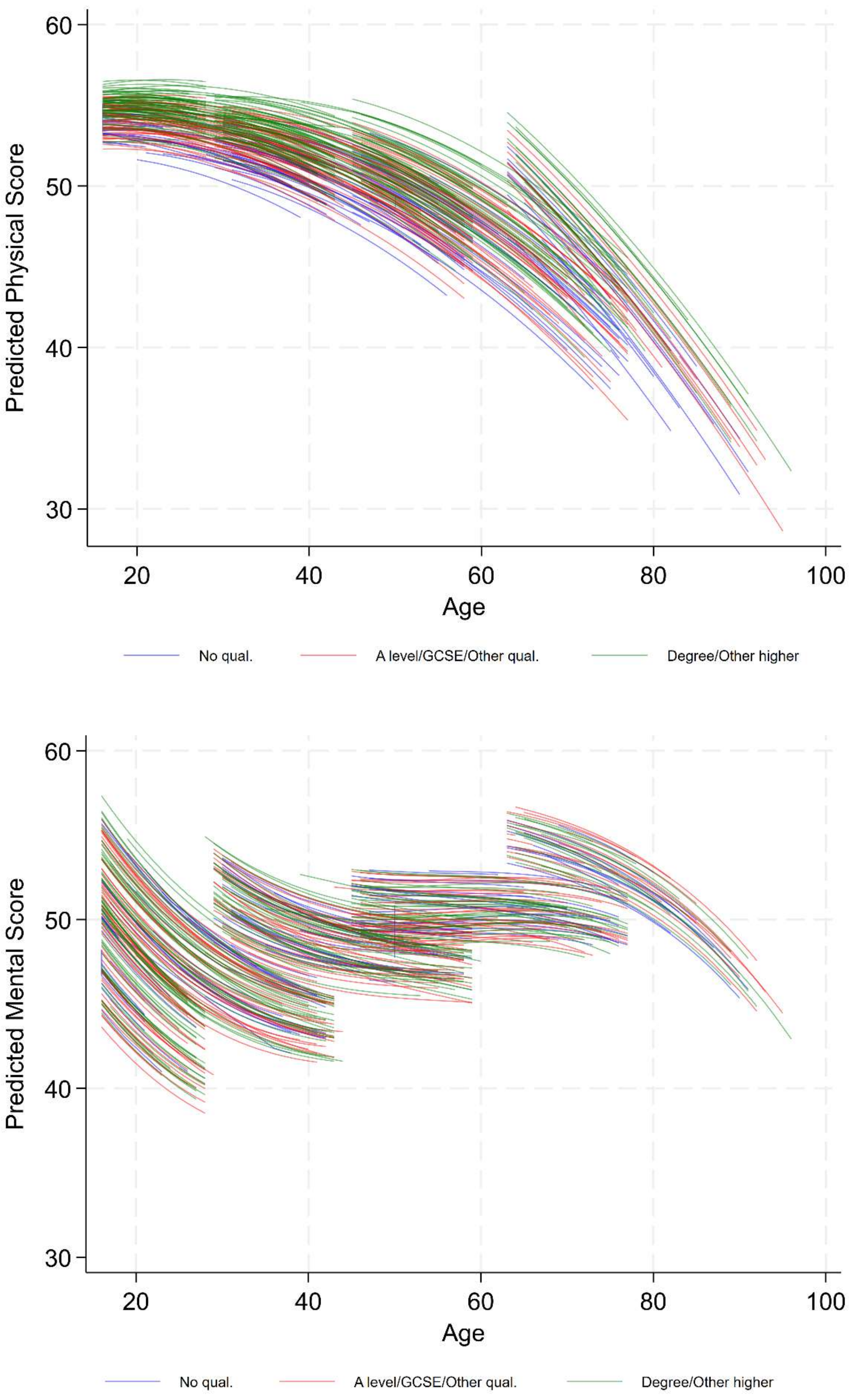

***Figure S6. Predicted physical and mental health from Model 2 coloured by socioeconomic class***

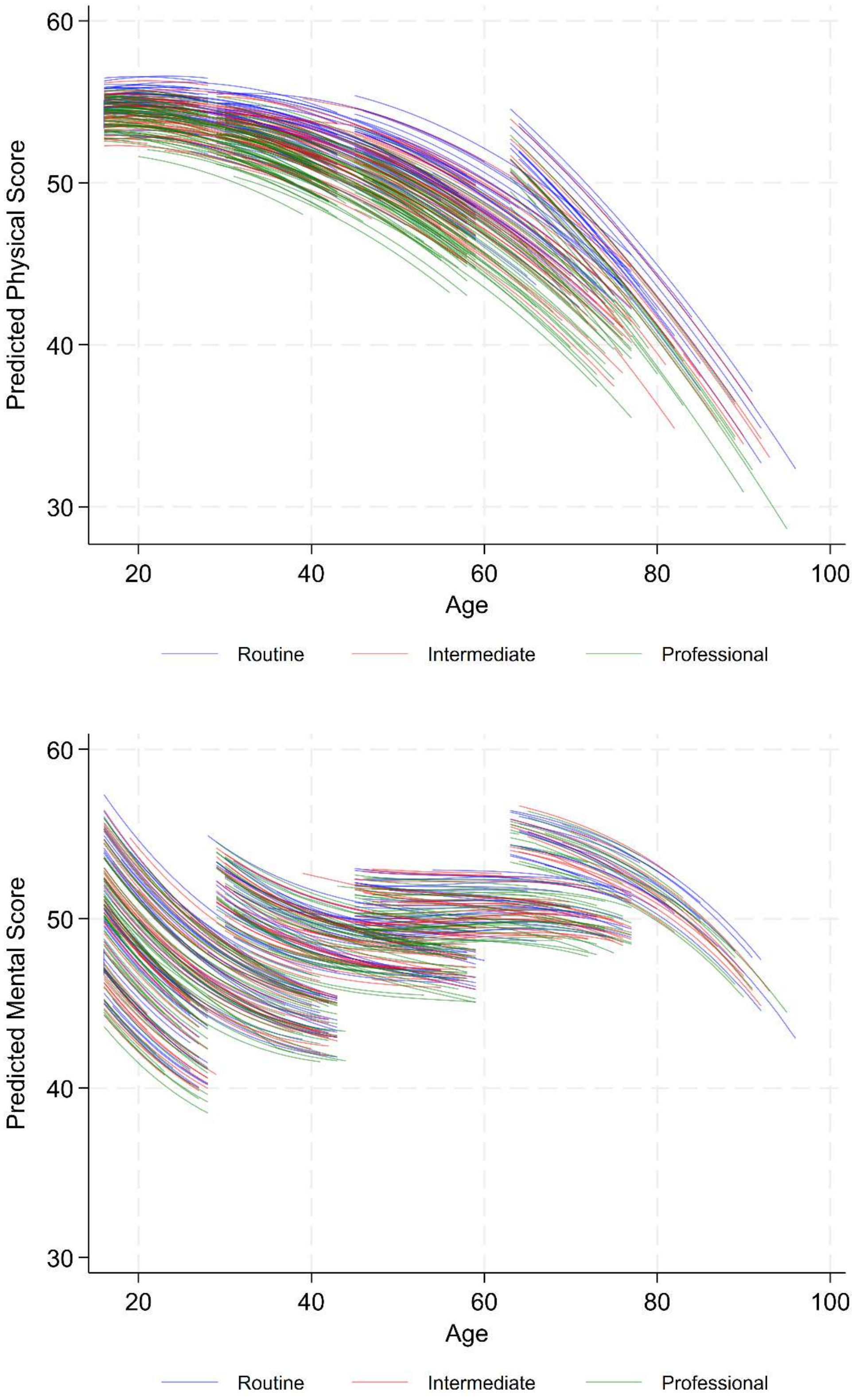

***Figure S7. Predicted mental health from Model 2 by sex and ethnicity for White and Black groupings***

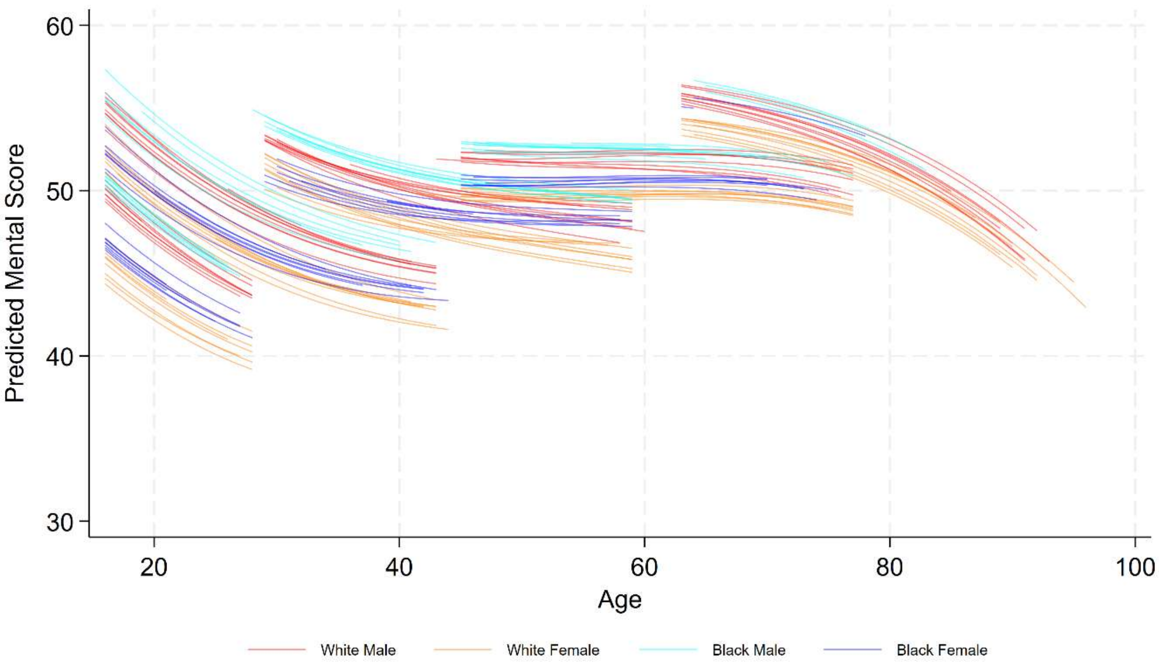

***Figure S8. Predicted mental health from Model 2 by sex and ethnicity by White and Asian groupings***

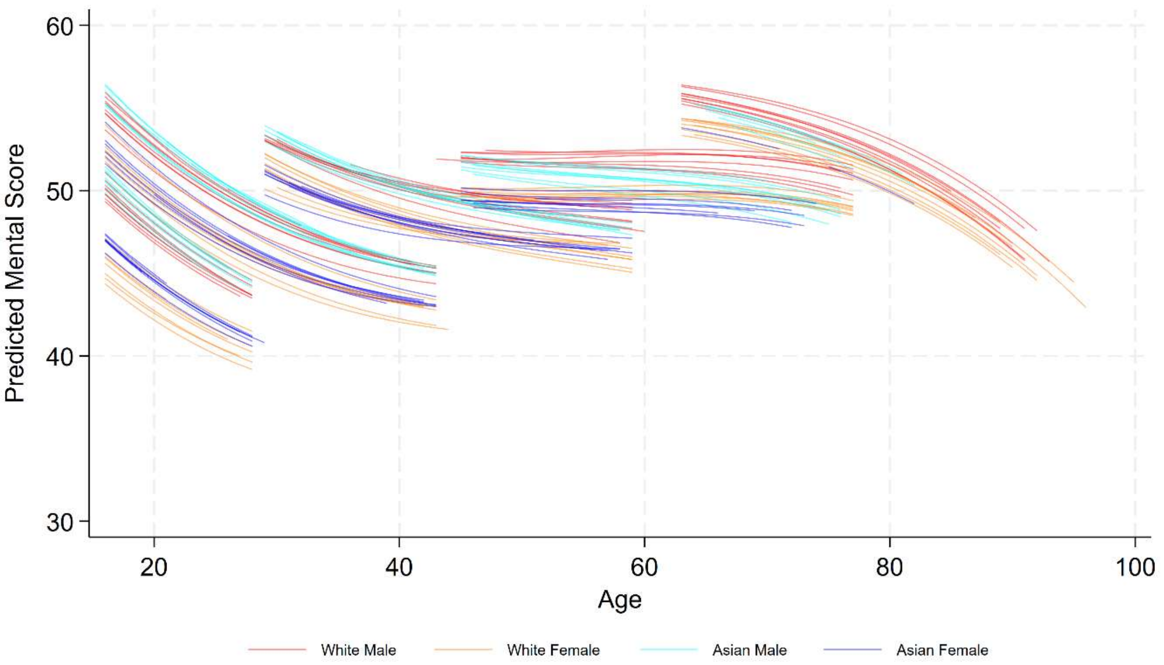

***Figure S9. Predicted mental health from Model 2 by sex and ethnicity by White and Mixed groupings***

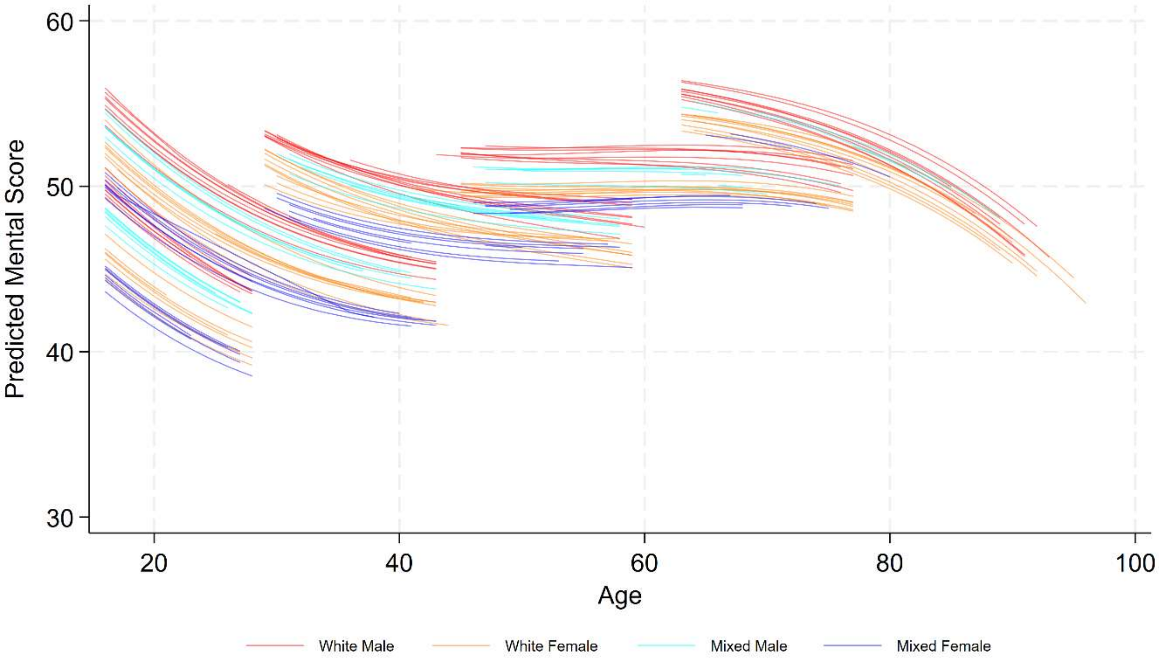

***Figure S10. Predicted mental health from Model 2 by sex and ethnicity by White and Other groupings***

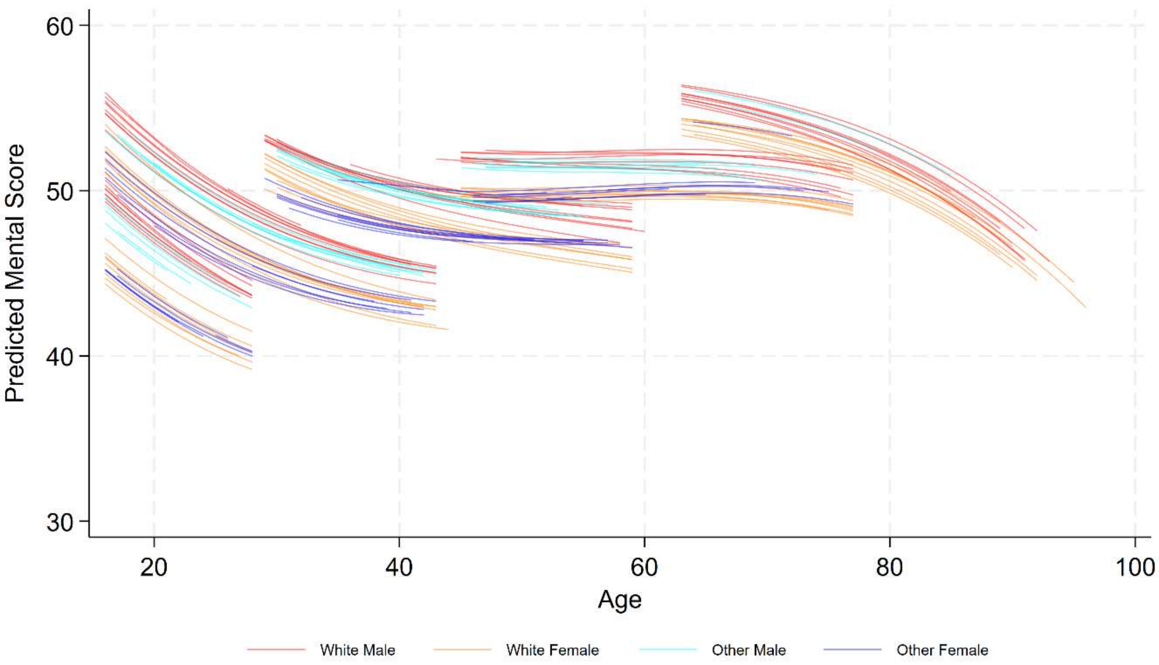

***Figure S11. Number of strata category combinations that are filled by our dataset shown by sex, generation and ethnicity.***

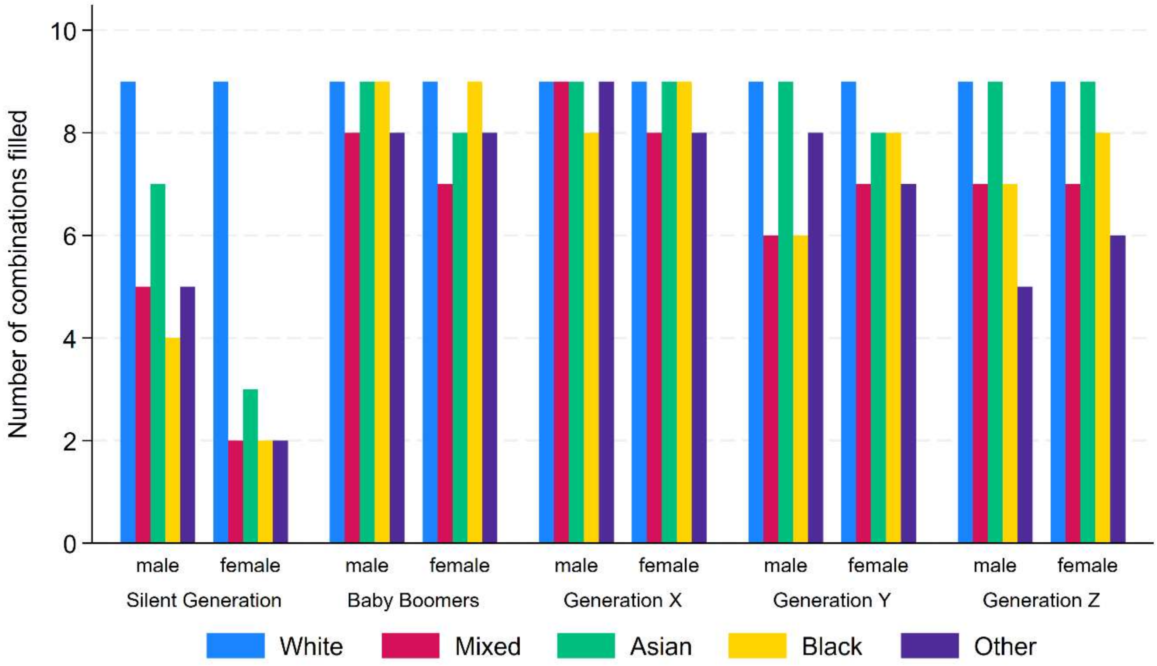


Note: Each sex by generation by ethnicity combination should have 9 strata, to account for the remaining highest qualification and socioeconomic class combinations (3 * 3).

***Figure S12. Scatterplots of the Model 1 physical and mental health intercepts where outlying strata by the difference between the two outcomes are highlighted in black.***

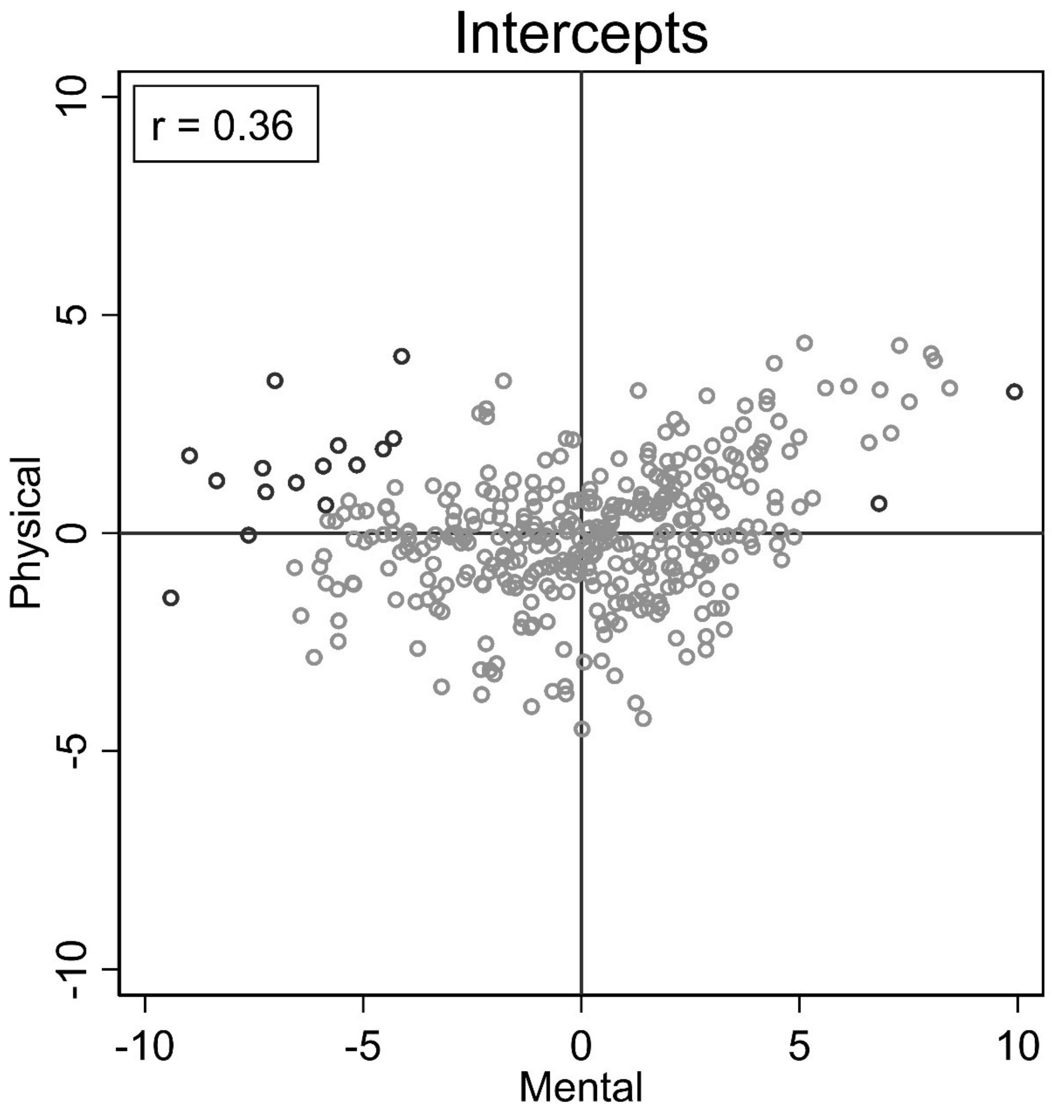
